# Minority and Rural Coronavirus Insights Study (MRCIS): The Need for Targeted SARS CoV-2 Vaccination Efforts in Minority Populations

**DOI:** 10.1101/2021.10.06.21264407

**Authors:** Latrice G. Landry, LaTasha Lee, Nishanth Chalasani, Liou Xu, Taylor Stair, Gary Puckrein, William A. Meyer, Gary Wiltz, Marian Sampson, Paul Gregerson, Charles Barron, Jeffrey Marable, Ola Akinboboye

**Author notes:** Corresponding Author Latrice G. Landry. Indicates equal contribution by authors (Latrice G. Landry, LaTasha Lee).

## Abstract

**Importance:** Racial and ethnic minority populations have been disproportionately affected in terms of hospitalizations and deaths during the COVID-19 pandemic. Vaccine uptake remains a barrier to full population inoculation against this highly infectious disease.

**Objective:** The purpose of this report is to describe SARS-CoV-2 vaccine interest rates in a racially, geographically, and ethnically diverse study cohort and characterize vaccine interest across a racially, ethnically, and geographically diverse study population.

**Design:** This report describes responses to a survey administered between November 2020 and May 2021 using a community convenience sample through a partnership between the National Minority Quality Forum (NMQF) and Federally Qualified Health Centers (FQHCs) as part of the Minority and Rural Coronavirus Insights Study (MRCIS). Analysis of survey responses from 3,624 participants are provided.

**Results:** Early data from the MRCIS cohort suggest that “SARS-CoV-2 vaccine hesitancy” is more prevalent in Black versus Non-Hispanic Whites survey respondents, and the Hispanic community has positive interest in the vaccine, to a similar degree as Whites. The persistent presence of “vaccine undecideds” across different sites and racial/ethnic groups uncovers the need for more public health efforts to influence positive views about vaccination.

**Conclusion:** These findings highlights the urgent need for interventional educational campaigns targeted at populations at risk of low vaccine interest. Focused efforts are needed to combat misinformation and explain the vaccine’s safety and effectiveness to promote its uptake and avoid low inoculation rates. Public health communication must consider differences in population groups, regions, and social determinants of health to fully address vaccine uptake disparities and overcome alleged hesitancy.

**Key Points:**

- Willingness to receive the SARS CoV-2 varies among minority populations.
- “SARS-CoV-2 vaccine hesitancy” is more prevalent in the non-Hispanic Black population than the non-Hispanic White and Hispanic populations.
- Public health infrastructure is needed in underserved communities for efficient assessment and targeted communication of public health priorities such as the SARS CoV-2 vaccination.

## Introduction:The Disproportionate Impact of COVID-19 on Minority Populations

Since the onset of the COVID-19 (Coronavirus Disease 2019) pandemic, minority populations have been disproportionately affected in terms of hospitalizations and deaths. In the initial stages of the outbreak, as early as April 8, 2020, the Centers for Disease Control and Prevention (CDC) released data that examined approximately 1,500 patients hospitalized due to the COVID-19 across 14 different states. The agency found that Black Americans accounted for 33 percent of hospitalizations, even though they only comprise 18 percent of the U.S. population.^1^ July 2021 CDC data indicated that Black, non-Hispanic persons were 2.8 times more likely to be hospitalized for COVID-19 infection and nearly twice as likely to die from the virus when compared to White, non-Hispanic populations.^2^

Racial and ethnic minorities such as Black and Hispanic populations may be at greater risk for serious COVID-19 outcomes due to comorbidities such as diabetes, obesity, heart disease, and hypertension.^3^ In addition, racial and ethnic minorities are more likely to live in densely populated neighborhoods often in multigenerational households with lower socioeconomic status, leading to increased opportunities for virus transmission and limited access to care (including SARS-CoV-2 testing).^4^ The U.S. Dept of Labor statistics reported that in the years just prior to the pandemic, 19.7% of Black employees were able to work from home compared to 29.9% of White employees,^5^ thereby placing them at greater risk of infection through community exposure. This confluence of factors has placed the Black and Hispanic communities at the epicenter of the COVID-19 health care crisis. Consequently, there is a need to better understand why racial and ethnic minorities are disproportionately affected by COVID-19 and to continue to assess factors associated with interest in and access to SARS CoV-2 vaccines. These factors are contributing to the excess burden of death and disability from COVID-19 infections in these populations.

While there are reports suggesting lower SARS CoV-2 vaccination rates in minorities, these rates have changed over time.^6^ Recent reports have challenged the use of the term “vaccine hesitancy” in minority communities as vaccine interest may be closely linked with access to information^7^ and equitable vaccine distribution.^8^ As noted in a recent publication by Schoch-Spana et al., “Vaccine uptake, and especially the widespread acceptance of vaccines, is a social endeavor that requires consideration of human factors.”^9^ The variability of the human factors affecting minority interest in SARS CoV-2 vaccination is not well understood. Understanding inter/intra-variability within and between minority groups is essential as effective policies and interventions require nuanced understanding of the obstacles. However, despite a clear need for investing in understanding vaccine interest in minority communities, there has been limited research in this area.

### Early Vaccination Efforts

The U.S. Food and Drug Administration (FDA) authorized for emergency use the first two-dose SARS CoV-2 vaccines in December 2020 (Pfizer and Moderna). The first single-dose vaccine (Janssen by Johnson & Johnson) was authorized for emergency use in late February 2021 and was put on hold for a period following some adverse event reports. Initial vaccination uptake was reported to be lower among communities of color. Media reports suggested that “vaccine hesitancy” in these traditionally underserved communities combined with historical mistrust of the medical and research enterprises within the Black community might account for the lower vaccination uptake. This explanation overlooks some of the inherent biases in the phased rollout and disparate accessibility to vaccine distribution sites. For example, an early strategy to restrict doses to those age 75 and older ignored the reality that average life expectancy for the non-Hispanic Black population in 2020 was 72 years compared to 78 years for the non-Hispanic, White population.^10^ Moreover, in 2018, 9% of Black Americans were 65 and older and 8% of persons of Hispanic origin were in that age group, compared to 20% of non-Hispanic Whites.^11^ As such, it should not be surprising that the White population was being vaccinated at rates higher than that of minority populations. Kaiser Family Foundation data show that across 40 states as of May 24, 2021, the percent of the White population that had received at least one SARS CoV-2 vaccine dose (43%) was roughly 1.5 times higher than the rate for the Black population (29%) and 1.3 times higher than the rate for the Hispanic population (32%).^12^

As of July 29, 2021, CDC’s reported demographic characteristics, including race/ethnicity when available, of those receiving at least one dose of a SARS CoV-2 vaccine nationally showed the White, non-Hispanic population four times more likely than the Hispanic population and six times more likely than the Black, non-Hispanic population to have met that milestone.^13^

The purpose of this report is to describe the current vaccine acceptance rates in a racially, geographically, and ethnically diverse study cohort and characterize vaccine interest across a racially, ethnically, and geographically diverse study population.

## Methods

In late 2020, the National Minority Quality Forum (NMQF) launched the Minority and Rural Coronavirus Insights Study (MRCIS), a prospective longitudinal investigation of risk and socioeconomic factors associated with the disproportionate impact of COVID-19 on minority and rural communities. Federally Qualified Health Centers (FQHCs), funded through the Health Resources & Services Administration (HRSA), were invited to be MRCIS partners because they are community-based health care providers, largely serving Medicaid and uninsured populations. In 2014, more than 25% of those living in poverty were seen by FQHCs compared to less than 1% of those with incomes greater than 200% of the Federal Poverty Level.^14^ FQHCs also serve higher proportions of patients in minority groups, particularly non-Hispanic Black and American Indian populations.^14^ Further, FQHCs were established to operate in communities that are underserved and therefore have been underrepresented in research. To date, five community health centers in four HRSA regions (4, 5, 6, and 9) and five states have been selected to participate in MRCIS (see Table 1). Research sites were selected based on the high proportion of deaths in minority populations that they serve in proportion to the proportion of minorities in the state population.

**Table 1.** Demographics of MRCIS Vaccine Survey Respondents by Site (as of May 31, 2021) (n=3624)

|  | **HRSA 6**<br>(Louisiana)<br>(n=1020) | **HRSA 4**<br>(Florida)<br>(n=1019) | **HRSA 5**<br>(Illinois and Ohio)<br>(n=583) | **HRSA 9**<br>(California)<br>(n=1002) |
| --- | --- | --- | --- | --- |
| **Mean age (SD)** | 57.1 (15.6) | 50.3 (16.0) | 42.8 (14.0) | 46.2 (13.9) |
| **Percent age 65 and older** | 34.20% | 21% | 7.1% | 8.9% |
| **Sex at Birth** |  |  |  |  |
| Male | 376 (37%) | 343 (34%) | 234 (40%) | 488 (49%) |
| Female | 643 (63%) | 674 (66%) | 339 (58%) | 514 (51%) |
| Other/Blank | 1 | 2 | 10 (2%) | 0 |
| **Race** |  |  |  |  |
| Black, non-Hispanic | 545 (53%) | 50 (5%) | 95 (16%) | 183 (18%) |
| White, non-Hispanic | 393 (39%) | 70 (7%) | 131 (22%) | 134 (13%) |
| Other | 34 (3%) | 38 (4%) | 94 (16%) | 70 (7%) |
| **Ethnicity** (identified as Hispanic or Latino) | 48 (5%) | 861 (84%) | 263 (45%) | 615 (61%) |

Initial recruitment of participants consisted of a community convenience sample of adults in minority communities at HRSA sites. The populations within these communities include non-Hispanic Black, Hispanic, American Indian, and non-Hispanic White populations. The five HRSA sites are located in urban, suburban, and rural populations, which are geographically dispersed on the West Coast, and Midwest, Southeast, and Southern parts of the United States. (Table 1) The protocol, which was reviewed and approved by an institutional review board (IRB), involved a phased-in approach of enrolling 5,000 adults over 5 years. The MRCIS study is designed to examine the impact of COVID-19 on participants and their household members.

MRCIS study participants were given viral and antibody testing for SARS CoV-2 in addition to completing a baseline survey indicating their race, ethnicity, age, gender, and sex. Participants were also asked questions regarding housing, utilities, broadband access, and basic needs. Additionally, participants were asked a series of questions regarding: any prior positive COVID-19 test results for themselves, members of their household, and friends and neighbors; and social distancing and mask wearing behaviors. Questions about vaccine interest focused on the SARS CoV-2 and flu vaccines. For the SARS CoV-2 vaccine, participants were asked “Would you get a coronavirus vaccine,” with the options to answer, “yes,” “no,” or “undecided.” Participants were also asked whether they would get a flu shot this year (“yes” or “no”). The first responses were recorded in November 2020 (before any vaccine was available) and continued through April 2021.The SARS CoV-2 vaccine interest percentage was calculated for the sites within the four HRSA regions that the clinical research sites represent. Participant responses were stratified by race and by Hispanic ethnicity.

To summarize the assembled data, frequencies N (%) of the responses to the vaccine interest question were reported for all sites. The race/ethnicity variable was combined from the two separate variables of Race (White, Black, and Other), and Ethnicity (Hispanic, non-Hispanic, and Prefer Not to Answer), resulting in four mutually exclusive categories: White non-Hispanic, Black non-Hispanic, Hispanic, and Other/Unknown. The non-Hispanic White category served as a reference group in the pairwise comparisons. Pearson’s Chi-Square tests were used for comparisons by site and by race/ethnicity group. P-values were computed to evaluate differences on the acceptance rate (% yes), the rejection rate (% no), and the hesitancy rate (% undecided). The one-way frequency test was performed on the overall vaccine interest responses to test for equality of the proportions in each category.

## Results

We report here on preliminary data on vaccine interest by race/ethnicity and by HRSA region. As of May 31, 2021, a total of 3624 participants had been enrolled at the five sites, with the earliest enrollment beginning in mid-November 2020 at the Louisiana site in HRSA region 6. Sites varied in racial and ethnic composition of participants, with a high proportion of Black participants in Louisiana (55%) and high proportions of Hispanic participants in Florida (HRSA region 4; 84%) and California (HRSA region 9; 61%). (Table 1) HRSA region 6 (Louisiana) had the highest mean age (57 years), while region 5 had the lowest mean age (42.8 years). The highest percent male participation was in HRSA region 9 at 49% (California), while region 5 (Florida) had the lowest participation from men at 34%.

Reported housing insecurity was low (3-5%) in participants in HRSA regions 4, 5, and 6, but higher in region 9 (25%). Inability to pay gas, water, or electric was 3% in region 6, 6% in region 4, 7% in region 9, and 11% in region 5. Lack of access to broadband was 15% in region 4, 20% in region 6, 25% in region 5, and 28% in region 9. (Table 2) In region 9, 25.3% of participants identified as homeless. Of those who identified as homeless in this region, 68% were male, while 21%, 43%, and 36% identified as non-Hispanic White, non-Hispanic Black, and Hispanic ethnicity, respectively. (Table 2)

**Table 2.**
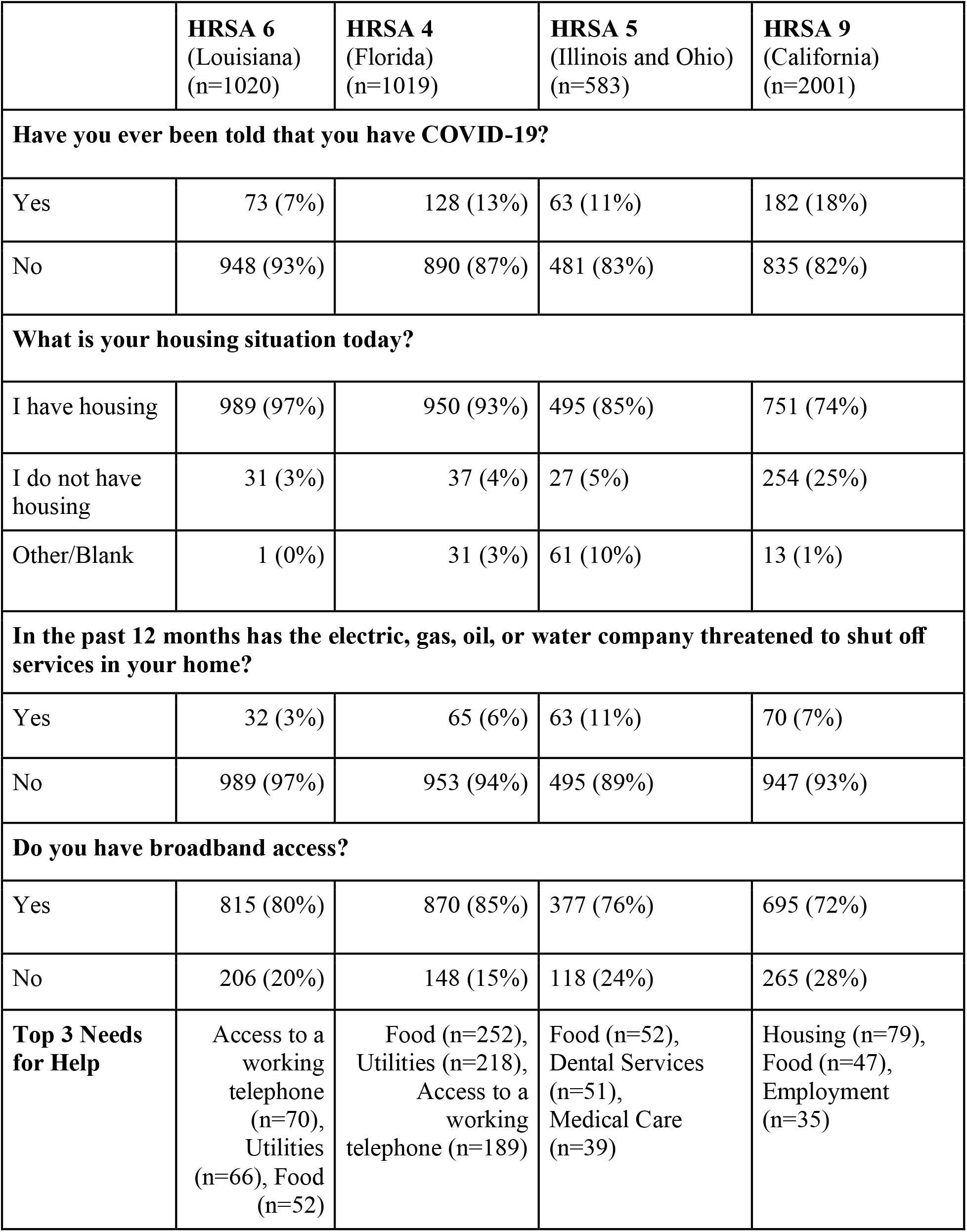
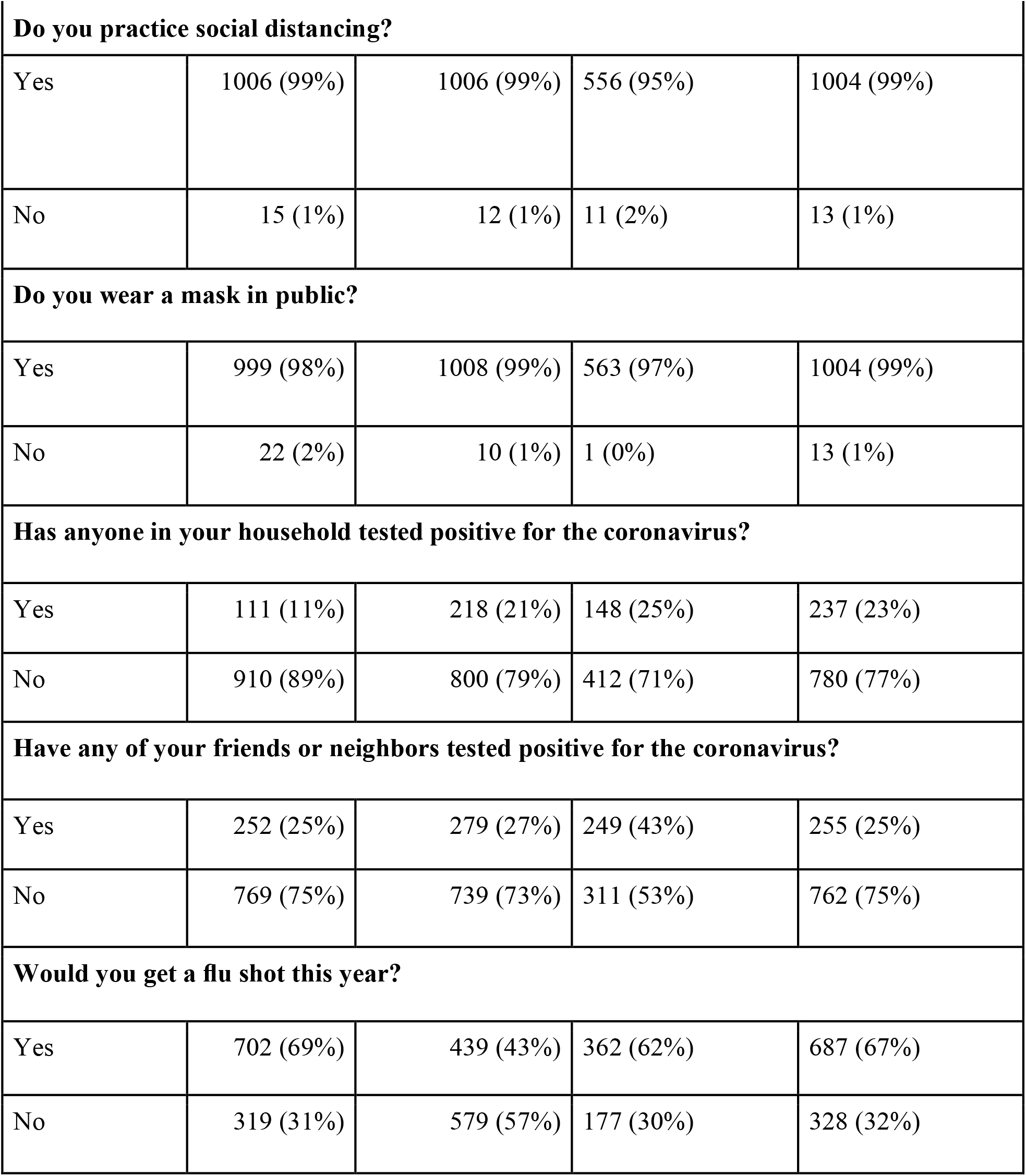
Select Socioeconomic Factors and Health Behaviors of Survey Respondents (as of May 31, 2021) (n=3624)

|  | **HRSA 6**<br>(Louisiana)<br>(n=1020) | **HRSA 4**<br>(Florida)<br>(n=1019) | **HRSA 5**<br>(Illinois and Ohio)<br>(n=583) | **HRSA 9**<br>(California)<br>(n=2001) |
| --- | --- | --- | --- | --- |
| **Have you ever been told that you have COVID-19?** |  |  |  |  |
| Yes | 73 (7%) | 128 (13%) | 63 (11%) | 182 (18%) |
| No | 948 (93%) | 890 (87%) | 481 (83%) | 835 (82%) |
| **What is your housing situation today?** |  |  |  |  |
| I have housing | 989 (97%) | 950 (93%) | 495 (85%) | 751 (74%) |
| I do not have housing | 31 (3%) | 37 (4%) | 27 (5%) | 254 (25%) |
| Other/Blank | 1 (0%) | 31 (3%) | 61 (10%) | 13 (1%) |
| **In the past 12 months has the electric, gas, oil, or water company threatened to shut off services in your home?** |  |  |  |  |
| Yes | 32 (3%) | 65 (6%) | 63 (11%) | 70 (7%) |
| No | 989 (97%) | 953 (94%) | 495 (89%) | 947 (93%) |
| **Do you have broadband access?** |  |  |  |  |
| Yes | 815 (80%) | 870 (85%) | 377 (76%) | 695 (72%) |
| No | 206 (20%) | 148 (15%) | 118 (24%) | 265 (28%) |
| **Top 3 Needs for Help** | Access to a working telephone (n=70), Utilities (n=66), Food (n=52) | Food (n=252), Utilities (n=218), Access to a working telephone (n=189) | Food (n=52), Dental Services (n=51), Medical Care (n=39) | Housing (n=79), Food (n=47), Employment (n=35) |
| **Do you practice social distancing?** |  |  |  |  |
| Yes | 1006 (99%) | 1006 (99%) | 556 (95%) | 1004 (99%) |
| No | 15 (1%) | 12 (1%) | 11 (2%) | 13 (1%) |
| **Do you wear a mask in public?** |  |  |  |  |
| Yes | 999 (98%) | 1008 (99%) | 563 (97%) | 1004 (99%) |
| No | 22 (2%) | 10 (1%) | 1 (0%) | 13 (1%) |
| **Has anyone in your household tested positive for the coronavirus?** |  |  |  |  |
| Yes | 111 (11%) | 218 (21%) | 148 (25%) | 237 (23%) |
| No | 910 (89%) | 800 (79%) | 412 (71%) | 780 (77%) |
| **Have any of your friends or neighbors tested positive for the coronavirus?** |  |  |  |  |
| Yes | 252 (25%) | 279 (27%) | 249 (43%) | 255 (25%) |
| No | 769 (75%) | 739 (73%) | 311 (53%) | 762 (75%) |
| **Would you get a flu shot this year?** |  |  |  |  |
| Yes | 702 (69%) | 439 (43%) | 362 (62%) | 687 (67%) |
| No | 319 (31%) | 579 (57%) | 177 (30%) | 328 (32%) |

All sites reported low individual rates of an existing COVID-19 diagnosis (7-18%), low to moderate rates of a COVID-19 diagnosis by a household member (11-25%), and moderate rates of a COVID-19 diagnosis in a friend or neighbor (22-43%). Additionally, study sites in all 4 HRSA regions reported high rates of mask wearing (97-99%) and social distancing (95-99%). When asked whether they would get a flu shot this year, a majority of participants in regions 5, 6, and 9 replied “yes” (59%, 62”, and “67% respectively). In contrast, 43% of participants from region 4 responded “yes”(Table 2).

Across all sites and populations, willingness to receive the SARS CoV-2 vaccine was 56%, with 25% selecting undecided, but responses varied by site and race. Categorized participant responses by site are presented in Figure 1. HRSA region 9 (California) had the highest reported interest in the vaccine (72%), followed by region 5 (69%), region 4 (49%), and region 6 (41%). Additionally, intention to receive the flu vaccine was the only question that differed with interest in the SARS CoV-2 vaccine, with more than 70% of those interested in receiving the SARS CoV-2 vaccine also planning to get flu vaccine, and under 40% of those not interested in the SARS CoV-2 vaccine planning to receive the flu vaccine (data not shown). In all regions, men showed more interest in SARS CoV-2 vaccination than women, with region 6 (Louisiana) showing the largest difference; 54% (men) compared to 33% (women). Overall, 40% of respondents at the Louisiana site expressed a willingness to be vaccinated with the SARS-CoV-2 vaccine and another 40% were undecided. This was the highest number of undecided participants of all sites. However, enrollment started and ended at that site mid-December 2020, which was before a vaccine was widely available. Of note, that site’s study population has the highest proportion of Black participants compared to other sites (Table 1). Additionally, HRSA region 9 had the latest calendar enrollment, beginning enrollment in 2021 after SARS CoV-2 vaccinations had been made available through an emergency use authorization.

Figure 2 compares vaccine perceptions by race and ethnicity across all sites. Non-Hispanic Black participants were least likely to say yes to getting a SARS-CoV-2 vaccine, with 45% saying yes compared to 56% of Whites and 60% of Hispanics. Participant numbers for “other” groups were small and have been combined. One of the more striking results is the rather large discordance in “yes” responses between Blacks and Whites and Hispanics in the California population, an approximately 20% difference (83% of non-Hispanic whites, 75% of Hispanics, and 55% of non-Hispanic Blacks interested). In the non-Hispanic Black group in this region, 30% were not interested in the vaccine, with only 16% undecided. However, in regions 4 and 6, which began recruitment earlier, a higher percent of non-Hispanic Blacks (34 and 40%, respectively) reported they were “undecided.” This trend of a higher percent of the population being “undecided” in the earlier cohorts (regions 4 and 6) was consistent across all racial and ethnic groups.

The p-values of the Pearson’s Chi-Square tests (<.0001) provided evidence of statistically significant differences on the vaccine acceptance rate, the rejection rate, and the hesitancy rate among the four sites. The sites in HRSA regions 5 and 9 had much higher vaccine acceptance rates than the sites in HRSA regions 4 and 6. Across the race/ethnicity groups, the most significant differences were due to higher vaccine acceptance rate and lower rejection rate in the White non-Hispanic population compared to that in the Black non-Hispanic population. Overall, the proportions (% yes, % no, and % undecided) were not equal. This should not be unexpected given the overall vaccine acceptance rate was above 50%.

## Discussion

These early data from the MRCIS cohort suggest that lack of interest in the SARS-CoV-2 vaccine is more prevalent in the non-Hispanic Black community than in the non-Hispanic White and Hispanic study populations. Our sample shows that vaccine interest in the Hispanic community is comparable to that of non-Hispanic Whites. According to a recent Kaiser Family Foundation survey, among those who have not yet been vaccinated, Hispanic adults were found to be about twice as likely as White adults to say they want to get a SARS CoV-2 vaccine as soon as possible but that access and concerns about eligibility and immigration status are barriers.^15^ The persistent presence of “vaccine undecideds” across different sites and racial/ethnic groups uncover the need for more public health efforts to foster positive perceptions of vaccination. This subpopulation, with those “undecided” in particular, stands to be the most influenced by positive public health outreach campaigns.

HRSA region 9 had the highest interest in SARS CoV-2 vaccination in comparison to all other sites, with HRSA region 6 showing the lowest. However, as previously stated, these two sites recruited individuals at different times during the vaccine development and roll out with HRSA region 6 recruiting and wrapping up prior to the emergency use authorization of the first available vaccine. In contrast, the HRSA region 9 site enrolled most participants in the January/February timeframe at the height of the pandemic in California and the point where fear and anxiety were highest. Hospitals were overcrowded and death rates were increasing.

The results show a striking discordance in “yes” responses between racial/ethnic groups in the California population (HRSA 9) regarding vaccine interest (an approximate 20% difference). It is important to note that this site was the only site to target outreach to homeless populations. Of the non-Hispanic Black participants at this site, 57% identified as homeless. Therefore this finding is likely impacted by homelessness and housing insecurity in this population. Interestingly, a portion of the homeless population was recruited through a shelter with strong ties to a community health organization. The relatively small number of non-Hispanic Blacks at that shelter showed higher interest in the SARS-CoV-2 vaccine, which may be associated with positive programming at that site. This might support the use of local grassroots approach with trusted community partners to inform and influence vaccine interest.

Our study included rural, urban, sub-urban populations in five separate states in four geographically defined HRSA regions. The diversity of the study population, which spans a variety of categories of those who are underrepresented in research, provides a valuable resource for diverse, uncaptured insights into the pandemic. While polling and vaccination records highlight minority groups receiving SARS CoV-2 vaccination at lower rates, our study suggests the importance of de-aggregating these groups to provide clarity in need and targeted solutions. While states and municipalities have shown great creativity to increase vaccination rates through lottery and other marketing strategies, our study suggests that many participants were more interested in basic needs such as food, housing, employment, and medical care. It is possible that the trusted local partners already working to provide those needs in communities could be valuable partners to inform, influence, and influence SARS CoV-2 vaccine uptake among the unvaccinated. The only item directly related to SARS CoV-2 interest in our study was interest in influenza vaccination, reflecting generally parallel levels of interest. This may suggest that meaningful collaboration between influenza and SARS CoV-2 vaccination campaigns is warranted.

MRCIS investigators intend to follow up with those participants who responded “no” to receiving the vaccine to understand the reasons for that response. And for populations recruited before a vaccine was available, follow up will seek information on whether perceptions have changed, and if so, what changed them? The responses might point to community or facility practices, policies, or perceptions that can be the focus of future more tailored public health strategies and policies to enhance vaccination rates. These early data demonstrate that minorities are not monolithic and the differences in population groups, regions, and social determinants of health likely play a critical role in an individual’s response to a public health pandemic. Additionally, in follow up with study participants investigators plan to examine vaccine uptake in the communities and convene focus groups or community advisory boards to understand barriers and challenges to vaccine acceptance locally in these diverse communities.

In the future, genomic, proteomic, and metabolomic assessments are planned as part of the MRCIS project. All participants received a SARS CoV-2 nucleic acid amplification test and antibody test at study initiation. Additional laboratory analyses (such as HbA1c, glucose, serum Vitamin D) are obtained at enrollment and especially critically abnormal values are provided to the site staff for appropriate additional clinical follow-up and/or clinical management of participants. Future research reports from this study will focus on metabolic, viral detection, comorbidity, and social determinants of health data over the 5 years of the project.

In sum, suppositions about the explanation for race/ethnic disparity and alleged vaccine hesitancy in minority communities should be critically evaluated. Further, mitigating racial disparities in the uptake of SARS CoV-2 vaccines is critical to reduce the disproportionate impacts of this virus on minority communities and to slow widening health disparities in the future. These early results are reminders that infrastructure needs to be place in underserved communities to introduce critical public health measures such as vaccines, recognizing the variability among needs and populations.

## Data Availability

All data reported in this study are held by the National Minority Quality Forum and are not consented for public sharing or dissemination. All data inquiries should be referred to the National Minority Quality Forum 1201 15th Street, NW, Suite 340, Washington, DC 20005.

## Acknowledgments

The National Minority Quality Forum was responsible for the design and conduct of the study; collection, management, analysis, and interpretation of the data; preparation, review, approval and the decision to submit the manuscript for publication. Quest Diagnostics provided in-kind and staffing support for the diagnostic tests during the enrollment process. The Centene Corporation provided startup support for the study. The authors acknowledge Kathi E. Hanna for her editorial assistance in preparing this manuscript.

## Notes

### Competing Interest Statement

The authors have declared no competing interest.

### Clinical Trial

This manuscript provides a cross-sectional analysis of the minority and Rural Coronavirus Insights Study and is not a registered clinical trial.

### Funding Statement

This research was funded in by the National Minority Quality Forum, Quest Diagnostics and the Centene Corporation as a part of a collaborative response to coronavirus impact in minority and rural communities.

### Author Declarations

The protocol, which was reviewed and approved by an institutional review board (IRB) at WCG IRB Puyallup, WA.

